# GLP-1 Receptor Agonists and Acute Diabetes Complications in Adults with Type 1 Diabetes: A Target Trial Emulation

**DOI:** 10.1101/2025.11.10.25339908

**Authors:** Hao Dai, Yao An Lee, Rotana Radwan, Amy J. Sheer, Tamara S. Hannon, Matthew R. Hayes, Ramon C. Sun, Jiang Bian, Jingchuan Guo

**Affiliations:** Department of Biostatistics & Health Data Science, Indiana University School of Medicine, Indianapolis, Indiana, USA; Department of Pharmaceutical Outcomes and Policy, University of Florida, College of Pharmacy, Gainesville, FL, USA; Department of Medicine, University of Florida College of Medicine, Gainesville, FL, USA; Department of Pediatrics, Indiana University School of Medicine, Indianapolis, Indiana, USA; Department of Psychiatry, Perelman School of Medicine, University of Pennsylvania, Philadelphia, PA, USA; Department of Biochemistry and Molecular Biology, University of Florida, Gainesville, FL, USA; Center for Advanced Spatial Biomolecule Research, University of Florida, Gainesville, FL, USA; Regenstrief Institute, Indianapolis, Indiana, USA

## Abstract

**Background:** To evaluate the association between Glucagon-like peptide-1 receptor agonists (GLP-1RA) use and the risk of acute diabetes complications among adults with type 1 diabetes (T1D) who were eligible for anti-obesity medication (AOM) treatment.

**Methods:** We employed a target trial emulation using EHR data from the OneFlorida+ network (2014–2024) to investigate the association between GLP-1RA initiation and acute diabetes complications among adults with T1D. Eligible participants were adults with a diagnosis of T1D and who met clinical criteria for AOM treatment. GLP-1RA initiators were 1:1 matched to non-initiators using time-conditional propensity scores. The primary outcome was the occurrence of diabetic ketoacidosis (DKA). Secondary outcomes included severe hypoglycemia, all-cause hospitalizations, and emergency department (ED) visits. Cox proportional hazards models were utilized to estimate hazard ratios (HRs) and 95% confidence intervals (CIs). We applied a causal learning approach to explore heterogeneous treatment effects.

**Findings:** The matched cohort included 651 GLP-1RA users and 651 non-users. For GLP-1RA users and non-users, the incidence rates were 13.5 vs. 21.8 per 1,000 person-years for DKA. Compared to non-users, GLP-1RA use was not significantly associated with incidence of DKA (HR 0.62 [95%CI 0.33-1.17]) or severe hypoglycemia (HR 0.52 [95%CI 0.17-1.55]); notably, GLP-1RA use was significantly associated with fewer hospitalizations (HR 0.74 [95%CI 0.62-0.90]) and ED visits (HR 0.73 [95%CI 0.57-0.92]).

**Interpretation:** Among adults with T1D and obesity, GLP-1RA use was not associated with an increased risk of DKA or severe hypoglycemia but was linked to fewer ED visits and hospitalizations.

**Funding:** The study was supported by National Institute of Diabetes and Digestive and Kidney Diseases (NIH/NIDDK) **R01DK133465.**

## Introduction

Type 1 diabetes (T1D) represents approximately 5% to 10% of all diabetes cases in the United States (U.S.).^1^ Individuals with T1D continue to face challenges avoiding acute complications such as severe hypoglycemia^2,^ and diabetic ketoacidosis (DKA).^3^ Severe hypoglycemia, characterized by dangerously low blood glucose levels, can lead to confusion, seizures, loss of consciousness, or even death if not promptly treated.^2^ DKA, classically resulting from insulin deficiency, leads to the accumulation of ketones in the blood, causing metabolic acidosis, dehydration, and, if untreated, can progress to severe pain, coma, and/or death.^3^ About 4.2% of individuals with T1D experience at least one hypoglycemia-related hospitalization annually,^4^ and DKA accounts for 61.6 cases per 10,000 diabetes-related hospitalizations in the U.S.^5^ Although T1D is traditionally considered a disease of lean people, overweight and obesity are becoming increasingly more common in individuals with T1D.^6,7^

Glucagon-like peptide-1 receptor agonists (GLP-1 RAs) are a well-established class of antihyperglycemic agents approved for T2D. They enhance glucose-dependent insulin secretion, suppress glucagon, and delay gastric emptying, which collectively improve glycemic regulation.^8,9^ Although GLP-1 RAs are associated with favorable outcomes in T2D and generally well tolerated aside from gastrointestinal adverse events, their use in T1D remains off-label and largely under-investigated. Still, they are increasingly being explored as adjunctive therapy in T1D to reduce insulin requirements and glycemic variability.^10,11^ However, their potential effects on hypoglycemia and DKA risk remain unclear.

GLP-1 RAs are thought to carry a low intrinsic risk of hypoglycemia due to their glucose-dependent action.^8,9^ However, hypoglycemia may still occur, especially when combined with insulin.^12^ Emerging evidence also indicates that some GLP-1 RAs may increase hypoglycemia risk even when not used with other glucose-lowering therapies.^13,14^ A network meta-analysis of 34 T2D trials (excluding insulin and sulfonylureas) found increased hypoglycemia risk with all GLP-1 RAs except albiglutide, with odds ratios ranging from 1.59 to 3.74.^13^ Real-world post-marketing data from the FDA Adverse Event Reporting System (FAERS) also showed hypoglycemia signals across six GLP-1 RAs (excluding albiglutide due to a paucity of data), including severe events. Most reports involved T2D, but some occurred in T1D patients as well.^14^ In parallel, concerns have emerged regarding the potential association between GLP-1 RAs and DKA. In a FAERS-based pharmacovigilance study, most DKA reports linked to GLP-1 RAs involved individuals with T2D. However, the signal disappeared when GLP-1 RAs were used with insulin and was stronger when insulin was absent, indicating that reduced or withheld insulin may contribute to the observed risk.^15^ Additionally, case reports have documented euglycemic DKA in individuals with T1D or latent autoimmune diabetes in adults following GLP-1 RA use, often triggered by gastrointestinal side effects (nausea, vomiting, and/or diarrhea) and reduced insulin intake; all collectively highlighting potential safety concerns when insulin is withheld or interrupted in patients with T1D.^16,17^

Given the expanding off-label use of GLP-1 RAs in T1D and the limited availability of real-world evidence, further evaluation on safety and potential benefit is warranted. This study aimed to examine the association between GLP-1 RA use and the risk of DKA among individuals with T1D using real-world data from OneFlorida+ network. To explore heterogeneous treatment effect (HTE), we incorporated a causal learning approach to further identify subgroups of patients who may derive the maximized benefit or experience potential harm from GLP-1RAs, and to uncover key moderators.

## Research Design and Methods

### Data Source

Data was from the OneFlorida+ clinical research network, which is part of the National Patient-Centered Clinical Research Network (PCORnet). The dataset spans from 2014 through 2024 and includes comprehensive, longitudinal electronic health records (EHRs) for approximately 21 million individuals across Florida, Georgia, and Alabama.

OneFlorida+ integrates data from 14 partnering healthcare systems and is formatted according to the PCORnet Common Data Model (CDM).^18^ The dataset, designated as a HIPAA-limited set, includes key temporal and geographic identifiers (e.g., dates and ZIP codes) and provides robust clinical and demographic information, including diagnoses, procedures, medication use, laboratory results, and vital signs.

### Study Design and Study Population

This study employed a retrospective cohort design with a new-user approach. An intention-to-treat analysis was conducted to examine the relationship between GLP-1RAs and the incidence of acute diabetes complications and the occurrence of hospitalization or ED visits in real world T1D patients who were eligible for anti-obesity medications (AOMs). This study followed the Strengthening the Reporting of Observational Studies in Epidemiology (STROBE) reporting guidelines (**eTable 1 in Supplement)** and was approved by the University of Florida Institutional Review Board (# **IRB202300903**).

The study cohort comprised T1D adults meeting the eligible criteria for anti-obesity medication (AOM) treatment. (1) According to 2013 AHA/ACC/TOS guideline, which is applied to now, AOM eligibility was defined as the presence of a documented diagnosis of obesity (BMI ≥ 30 kg/m²) or a BMI in the range of 27.0 to 29.9 kg/m² accompanied by at least one weight-related comorbidity.^19^ (2) Because of the high prevalence of T2D in the adult population, adult-onset T1D is misclassified as T2D in at least one in three cases in clinical practice.^20^ Simple ICD codes may not provide accurate identification of T1D in real-world data. Therefore, we adopted a validated EHR-based computable phenotype developed through network-wide chart reviews, which achieved >90% sensitivity, specificity, positive predictive value (PPV), and negative predictive value (NPV) in classifying T1D. ^21,22^ To further reduce bias, we additionally restricted the cohort to individuals with documented insulin use.

We then identified GLP-1RA users and non-users. The index date for GLP-1RA users was defined as the date of the first GLP-1RA prescription. The comparator group, defined as patients who did not initiate GLP-1RA therapy during the study period, was constructed by aligning non-users to GLP-1RA users within ±1 week window of the initiation date, to ensure temporal comparability of treatment assignment. Individuals were excluded if they were under 18 years of age at the index date, had not been identified as T1D before the index date, or lacked any clinical encounters in the 12 months preceding the index date. Patients’ follow-up began at the index date and continued until the earliest occurrence of one of the following censoring events: the outcome of interest, death, loss to follow-up (the date of the last recorded clinical encounter), or the end of the study period (January 31, 2024).

### Study Outcomes

The primary outcome was hospitalization or ED visit for DKA. The secondary outcomes included severe hypoglycemia (i.e., hospitalization or ED visit for hypoglycemia), any hospitalization, and ED visit. DKA and severe hypoglycemia were identified using ICD-9 and ICD-10 diagnosis codes, based on previously validated computable phenotype definitions^23–25^ (**eTable 2 in Supplement**).

### Exposure of interest

The exposure of interest was the new use of GLP-1RA, i.e., semaglutide, tirzepatide, and liraglutide, identified using RxNorm codes and National Drug Codes (NDCs) (**eTable 3 in Supplement**). The comparator group consisted of non-users of GLP-1RAs.

### Covariates

Baseline covariates encompassed a range of demographic, clinical, and pharmacological characteristics, as detailed in **Table 1**. Demographic variables included age, sex, race/ethnicity, smoking status, and insurance type. Clinical covariates comprised comorbid conditions (e.g., cardiovascular disease, biliary disorders and neuropathy), clinical observations (e.g., BMI, blood pressure), laboratory values (e.g., hemoglobin A1c [HbA1c], estimated glomerular filtration rate [eGFR], and lipid profile), and medication (e.g., aspirin, opioids, and statins).

**Table 1.** Baseline characteristics of GLP-1RA Users vs GLP-1RA Non-Users cohort after 1:1 propensity score matching.

| Characteristics | GLP-1RA Users, No.(%) (n=651) | GLP-1RA Non-Users, No.(%) (n=651) | SMD |
| --- | --- | --- | --- |
| Age, yrs, mean(std) | 44.57 (14.64) | 45.01 (17.44) | -0.0274 |
| **Race/Ethnicity** |  |  |  |
| Hispanics | 136 (20.89)% | 122 (18.74)% | 0.0539 |
| NHB | 126 (19.35)% | 126 (19.35)% | 0.0000 |
| NHW | 306 (47.0)% | 305 (46.85)% | 0.0031 |
| Others | 83 (12.75)% | 100 (15.06)% | 0.0384 |
| **Sex** |  |  |  |
| Female | 413 (63.44)% | 415 (63.75)% | -0.0064 |
| Male | 238 (36.56)% | 236 (36.25)% | 0.0064 |
| **Vital information** |  |  |  |
| BMI | 34.11 (6.21) | 34.02 (7.28) | 0.0120 |
| Systolic blood pressure | 129.83 (15.37) | 129.75 (16.13) | 0.0051 |
| Diastolic blood pressure | 77.61 (9.88) | 77.7 (10.07) | -0.0088 |
| **Insurance Coverage** |  |  |  |
| Medicaid | 35 (5.38)% | 48 (7.37)% | -0.0817 |
| Medicare | 67 (10.29)% | 72 (11.06)% | -0.0249 |
| No Payment | 15 (2.3)% | 14 (2.15)% | 0.0104 |
| Others | 76 (11.67)% | 91 (13.98)% | -0.0689 |
| Private | 205 (31.49)% | 191 (29.34)% | 0.0467 |
| Unknown | 253 (38.86)% | 235 (36.1)% | 0.0571 |
| **Smoking Status** |  |  |  |
| Current Smoker | n<11 | n<11 | -0.1300 |
| Former Smoker | n<11 | n<11 | 0.0554 |
| Never Smoker | 53 (8.14)% | 40 (6.14)% | 0.0775 |
| Unknown | 596 (91.55)% | 603 (92.63)% | -0.0398 |
| **Lab** |  |  |  |
| HbA1c | 8.28 (1.75) | 8.33 (2.16) | -0.0278 |
| HDL | 51.92 (11.97) | 52.45 (11.31) | -0.0456 |
| LDL | 88.09 (12.2) | 89.43 (13.06) | -0.1057 |
| Triglycerides | 138.78 (85.73) | 137.16 (83.54) | 0.0192 |
| Total cholesterol | 171.19 (33.89) | 172.1 (33.56) | -0.0269 |
| eGFR | 80.83 (23.14) | 80.85 (24.26) | -0.0009 |
| **Medications** |  |  |  |
| Insulin | 651 (100.0)% | 651 (100.0)% |  |
| ACEIs | 163 (25.04)% | 160 (24.58)% | 0.0108 |
| Beta-blocker | n<11 | n<11 | 0.0116 |
| CCB | n<11 | n<11 |  |
| Diuretic | 18 (2.76)% | 20 (3.07)% | -0.0098 |
| Angiotensin-receptor blocker | 118 (18.13)% | 113 (17.36)% | -0.0126 |
| Statin | 271 (41.63)% | 271 (41.63)% | -0.0319 |
| Lipid-lowering non-statin | 44 (6.76)% | 39 (5.99)% | 0.0314 |
| NSAIDS | 104 (15.98)% | 111 (17.05)% | -0.0289 |
| Warfarin | n<11 | n<11 | 0.0000 |
| Direct oral anticoagulant | 19 (2.92)% | 26 (3.99)% | -0.0588 |
| Aspirin | 83 (12.75)% | 84 (12.9)% | -0.0046 |
| Non-aspirin antiplatelet agents | 25 (3.84)% | 21 (3.23)% | 0.0333 |
| Proton pump inhibitor | 123 (18.89)% | 113 (17.36)% | 0.0399 |
| Antidepressant | 77 (11.83)% | 76 (11.67)% | 0.0048 |
| Antipsychotics | 43 (6.61)% | 43 (6.61)% | 0.0000 |
| Antiparkinson agent | 53 (8.14)% | 53 (8.14)% | 0.0000 |
| Benzodiazepines | 88 (13.52)% | 86 (13.21)% | 0.0090 |
| HRT | 20 (3.07)% | 17 (2.61)% | 0.0277 |
| Anti-dementia medication | n<11 | n<11 | 0.0210 |
| Oral steroids | 165 (25.35)% | 157 (24.12)% | 0.0285 |
| Opioid | 147 (22.58)% | 147 (22.58)% | 0.0000 |
| TNF inhibitor | n<11 | n<11 | 0.0000 |
| Immunosuppressants | 15 (2.3)% | 7 (1.08)% | 0.0954 |
| Orlistat | n<11 | n<11 |  |
| Phentermine topiramate | n<11 | n<11 | 0.0320 |
| Bupropion/Naltrexone | n<11 | n<11 | -0.0248 |
| Lorcaserin | n<11 | n<11 |  |
| **Weight-related comorbidities** |  |  |  |
| Cancer | 220 (33.79)% | 205 (31.49)% | 0.0491 |
| Acute/Chronic Pancreatitis | n<11 | n<11 | 0.0321 |
| Adenocarcinoma of the Esophagus | n<11 | n<11 |  |
| Anorexia | n<11 | n<11 | 0.0000 |
| Atrial fibrillation | 17 (2.61)% | 24 (3.69)% | -0.0616 |
| Breast cancers | n<11 | 12 (1.84)% | -0.0238 |
| Chronic kidney disease | 79 (12.14)% | 66 (10.14)% | 0.0635 |
| Colorectal cancers | n<11 | n<11 | -0.0185 |
| Coronary Artery Disease | 78 (11.98)% | 87 (13.36)% | -0.0415 |
| Cushing Syndrome | n<11 | n<11 | -0.0210 |
| Depression | 112 (17.2)% | 107 (16.44)% | 0.0205 |
| Dyspepsia | n<11 | n<11 | -0.0298 |
| Dyslipidemia | 401 (61.6)% | 416 (63.9)% | -0.0476 |
| Feeding Difficulties | 59 (9.06)% | 64 (9.83)% | -0.0262 |
| Gallbladder Disease | n<11 | n<11 | -0.0432 |
| GERD | 117 (17.97)% | 125 (19.2)% | -0.0316 |
| Heart Failure | 29 (4.45)% | 42 (6.45)% | -0.0880 |
| Hypertension | 378 (58.06)% | 377 (57.91)% | 0.0031 |
| Inflammatory Bowel Disease | n<11 | n<11 | 0.0554 |
| Kidney cancer | n<11 | n<11 | 0.0320 |
| Liver Cancer | n<11 | n<11 | 0.0320 |
| Meningioma | n<11 | n<11 | 0.0000 |
| Metabolic Syndrome | 29 (4.45)% | 16 (2.46)% | 0.1094 |
| Multiple myeloma | 192 (29.49)% | 182 (27.96)% | 0.0339 |
| NAFLD/NASH | 49 (7.53)% | 49 (7.53)% | 0.0000 |
| Ovaries cancer | n<11 | n<11 |  |
| Pancreatic cancer | n<11 | n<11 | 0.0000 |
| Prader-Willi Syndrome | n<11 | n<11 |  |
| Pulmonary Embolism | n<11 | n<11 | 0.0000 |
| Rheumatoid Arthritis/Osteoarthritis (RA/OA) | 126 (19.35)% | 145 (22.27)% | -0.0719 |
| Sleep apnea | 82 (12.6)% | 93 (14.29)% | -0.0495 |
| Stroke/TIA | 13 (2.0)% | 20 (3.07)% | -0.0684 |
| Thyroid cancer | n<11 | n<11 | 0.0210 |
| Upper stomach cancer | n<11 | n<11 |  |
| Uterus cancer | n<11 | n<11 | 0.0554 |
| Venous Thromboembolism | 17 (2.61)% | 13 (2.0)% | 0.0409 |
| Vitamin D deficiency | 209 (32.1)% | 195 (29.95)% | 0.0465 |
| At least one weight related comorbidities | 629 (96.62)% | 633 (97.24)% | -0.0356 |
| **Other Comorbidities** |  |  |  |
| AD | n<11 | n<11 | 0.0000 |
| ADRD | n<11 | n<11 | -0.0630 |
| Acute kidney injury | 72 (11.06)% | 70 (10.75)% | 0.0098 |
| Alcohol use disorder | n<11 | n<11 | 0.0248 |
| Anemia | 125 (19.2)% | 140 (21.51)% | -0.0572 |
| Anxiety | 158 (24.27)% | 161 (24.73)% | -0.0107 |
| Asthma | 53 (8.14)% | 61 (9.37)% | -0.0435 |
| B12_deficiency | n<11 | n<11 | -0.0210 |
| Benign prostatic hyperplasia | n<11 | n<11 | 0.0000 |
| Biliary disease | 17 (2.61)% | 22 (3.38)% | -0.0450 |
| Bipolar | 15 (2.3)% | 16 (2.46)% | -0.0101 |
| Cardiovascular disease | 90 (13.82)% | 101 (15.51)% | -0.0477 |
| Cataracts | 70 (10.75)% | 65 (9.98)% | 0.0252 |
| COPD | 38 (5.84)% | 43 (6.61)% | -0.0318 |
| Endometrial cancer | n<11 | n<11 | 0.0210 |
| Exercise | 13 (2.0)% | 16 (2.46)% | -0.0312 |
| Gastroparesis | 42 (6.45)% | 34 (5.22)% | 0.0524 |
| Glaucoma | 27 (4.15)% | 28 (4.3)% | -0.0076 |
| Gout | 13 (2.0)% | 18 (2.76)% | -0.0504 |
| Hearing impairment | 22 (3.38)% | 22 (3.38)% | 0.0000 |
| HHF | 29 (4.45)% | 42 (6.45)% | -0.0880 |
| Hip Pelvic Fracture | n<11 | n<11 | 0.0210 |
| Lower extremity ulcers | 48 (7.37)% | 50 (7.68)% | -0.0116 |
| Lung cancer | n<11 | n<11 | -0.0554 |
| Bladder cancer | n<11 | n<11 | 0.0554 |
| MCI | n<11 | n<11 | 0.0784 |
| Neuropathy | 433 (66.51)% | 435 (66.82)% | -0.0065 |
| Obstruction | n<11 | n<11 | -0.0556 |
| OCD | n<11 | n<11 | -0.0352 |
| Osteoporosis | 22 (3.38)% | 19 (2.92)% | 0.0264 |
| Parkinson | n<11 | n<11 | -0.0630 |
| Periodontitis | n<11 | n<11 | 0.0554 |
| Prostate cancer | n<11 | n<11 | 0.0907 |
| PTSD | 13 (2.0)% | 11 (1.69)% | 0.0228 |
| Schizophrenia | n<11 | n<11 | 0.0962 |
| Seizures | 14 (2.15)% | 11 (1.69)% | 0.0336 |
| Sleep disorder | 141 (21.66)% | 148 (22.73)% | -0.0259 |
| Substance use | 82 (12.6)% | 94 (14.44)% | -0.0539 |
| Suicide | n<11 | 12 (1.84)% | -0.0640 |
| TBI | n<11 | n<11 | 0.0557 |
| Hypothyroidism | 185 (28.42)% | 201 (30.88)% | -0.0538 |
| VD | n<11 | n<11 | -0.0554 |
| Vision impairment | n<11 | n<11 | 0.0298 |
| ESRD | 29 (4.45)% | 16 (2.46)% | 0.1094 |
| Bariatric procedure | 134 (20.58)% | 133 (20.43)% | 0.0038 |
| Bariatric surgery | 279 (42.86)% | 276 (42.4)% | 0.0093 |
| Pregnant | 36 (5.53)% | 40 (6.14)% | -0.0262 |
| **CCI Comorbidities** |  |  |  |
| Myocardial infarction | 199 (30.57)% | 190 (29.19)% | 0.0302 |
| Congestive heart failure | 34 (5.22)% | 45 (6.91)% | -0.0708 |
| Peripheral vascular disease | 40 (6.14)% | 44 (6.76)% | -0.0250 |
| Cerebrovascular disease | 29 (4.45)% | 37 (5.68)% | -0.0560 |
| Chronic pulmonary disease | n<11 | n<11 | -0.0630 |
| Rheumatic disease | 80 (12.29)% | 92 (14.13)% | -0.0544 |
| Peptic ulcer disease | 15 (2.3)% | 19 (2.92)% | -0.0385 |
| Mild liver disease | n<11 | n<11 | -0.0168 |
| Diabetes without chronic complication | 64 (9.83)% | 64 (9.83)% | 0.0000 |
| Diabetes with chronic complication | 637 (97.85)% | 640 (98.31)% | -0.0336 |
| Hemiplegia or paraplegia | 225 (34.56)% | 214 (32.87)% | 0.0357 |
| Renal disease | n<11 | n<11 | -0.0745 |
| Any malignancy, including lymphoma and leukemia, except malignant neoplasm of skin | 87 (13.36)% | 73 (11.21)% | 0.0655 |
| Moderate or severe liver disease | 34 (5.22)% | 20 (3.07)% | 0.1079 |
| Metastatic solid tumor | n<11 | n<11 | 0.0000 |
| AIDS/HIV | n<11 | n<11 | 0.0279 |
| CCI Score | 2.3 (2.08) | 2.16 (1.83) | 0.0198 |
| **Outcomes** |  |  |  |
| DKA | 16 (2.5%) | 24 (3.7%) |  |
| Hospitalizations | 193 (29.6%) | 237 (36.4%) |  |
| ED visits | 124 (19.0%) | 156 (24.0%) |  |

SMD, standardized mean difference; AD/DR, Alzheimer's Disease and Related Dementias; COPD, Chronic obstructive pulmonary disease; ACEI, angiotensin-converting-enzyme inhibitors; ARB, angiotensin II receptor blockers; NSAIDs, non-steroidal anti-inflammatory drugs; TNF inhibitors, tumor necrosis factor inhibitors; HRT, hormone replacement therapy; GERD, gastroesophageal reflux disease; NAFLD, nonalcoholic fatty liver disease; NASH, nonalcoholic steatohepatitis; HHF, hypertensive heart failure; MCI, mild cognitive impairment; OCD, Obsessive-compulsive disorder; PTSD, Post-traumatic stress disorder; TBI, traumatic brain injury; ESRD, end-stage renal disease; CCI, Carlson comorbidity index; Others: include American Indian or Alaska Native, Asian, Native Hawaiian or Other Pacific Islander, and individuals identifying as multiracial.

### Statistical Analysis

To address potential confounding and enhance comparability between two treatment groups, we implemented a time-dependent 1:1 propensity score (PS) matching strategy.^26^ Propensity scores were derived from a multivariable logistic regression model incorporating a comprehensive set of baseline covariates. Matching was performed using a nearest-neighbor (KNN) algorithm without replacement and a caliper of 0.2, ensuring that each GLP-1RA user was matched to a non-user with the closest propensity score within the specified threshold.^27^ Covariate balance between matched pairs was evaluated using standardized mean differences (SMDs), with values below 0.1 considered acceptable balance between GLP-1RA users and non-users.

Furthermore, Cohen’s d^28^ was used to assess the magnitude of difference in covariates before and after matching, providing additional assurance that confounding was adequately minimized in the final matched cohort used for outcome analyses. Additional details are provided in **eMethod 1**.

We calculated the incidence rate (IR) of study outcomes and employed Kaplan–Meier survival analysis to illustrate the cumulative incidence per 1000 person-years over time between the matched groups. Additionally, Cox proportional hazard regression models were used to estimate hazard ratios (HRs) with 95% confidence intervals (CIs) by comparing GLP-1RAs users vs. non-users.

### Identification of Heterogenous Treatment Effect (HTE) and Potential Moderators

We employed a doubly-robust meta-learner^29^ following the potential outcome framework. A logistic algorithm was used to estimate the PS for GLP-1RA use, while an XGBoost regression model^30^ predicted the outcomes. To ensure unbiased estimation, we applied a triple cross-fitting strategy and used bootstrapping for hyperparameter tuning within each round. Additional methodological details are provided in **eMethod 2**. We estimated absolute risk differences (RDs) with 95% CIs to assess the average exposure effect, with negative RDs indicating benefit and positive values indicating harm. We conducted HTE analysis based on the estimations to explore risk variation using an interpretable decision tree.^31^ We also identified potential moderators based on key decision nodes that distinguish subpopulations with heterogeneous effects.

To further assess the strength of interaction effects for candidate moderators, we fit Cox proportional hazards models that included interaction terms of the treatment variable and candidate moderators, with robust variance estimation applied.^32^

### Subgroup Analysis

Subgroup analyses were conducted for both prespecified factors and factors identified through a data-driven approach via HTE modeling. The prespecified subgroups included: age groups (<40, 40–64, and ≥65 years), sex (male vs. female), racial and ethnic categories (non-Hispanic White, non-Hispanic Black, Hispanic, and other groups), obesity severity (overweight vs. obesity), baseline Metformin (use vs. non-use), and individual GLP1-RA drugs (liraglutide, semaglutide, and tirzepatide).

The second set of subgroup analyses involved potential moderators identified through HTE modeling. We conducted subgroup analyses to assess whether the treatment effect of GLP-1RA varied meaningfully across distinct patient strata, thereby evaluating whether the identified moderators consistently modified the effect in real-world clinical subgroups.

## Results

The flowchart of patient selection is presented in **Figure 1**. The study population comprised 15,585 T1D patients eligible for AOM treatment in OneFlorida+ from 2014-2024, including 664 GLP-1RA user and 14,921 GLP-1RA non-users. After applying 1:1 PS matching (**eFigure 1 & 2 in Supplement**), the final cohort included 1,302 matched participants (651 GLP-1RA user vs. 651 non-user). After PS matching, baseline characteristics showed a balanced distribution of key covariates, as detailed in **eTable 4 of Supplement**. In the matched cohort (**Table1**), mean age was 44.8 (± 16) years, 63.6% were female, and 43.4% were non-Hispanic White. The mean BMI at baseline was 34.6 kg/m2. The two treatment groups showed similar distributions in baseline HbA1c levels, hypertension prevalence, and other clinical factors.

**Figure 1.**
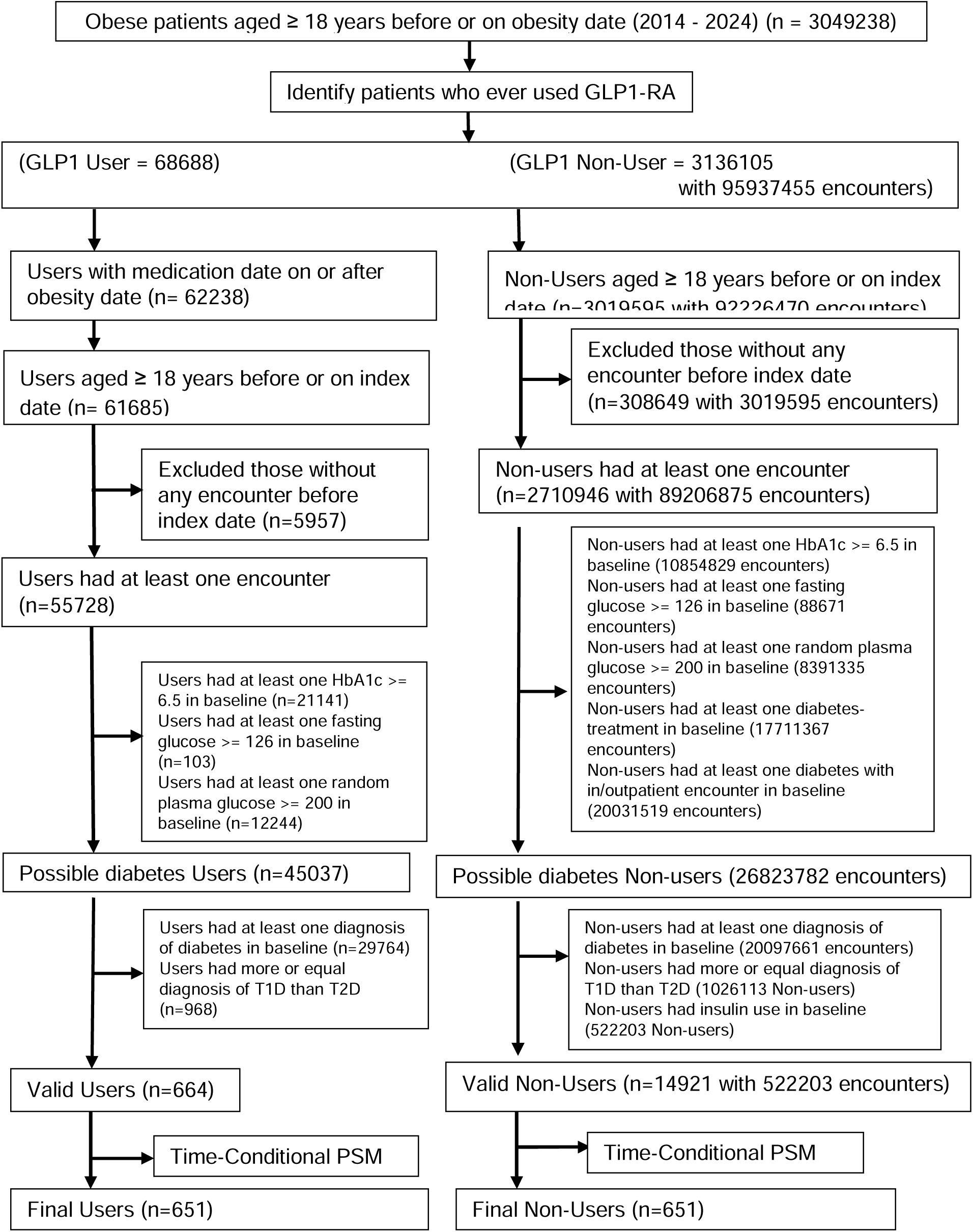
Flowchart of patient selection

eFigure 3 illustrates Kaplan-Meier plots depicting the cumulative incidence during follow-up. GLP-1RA users exhibited a lower incidence rate of DKA (13.5 per 1,000 person-years) compared with non-users (21.8 per 1,000 person-years). However, GLP-1RA use was not significantly associated with incidence of DKA (HR 0.62 [95%CI 0.33-1.17]) or severe hypoglycemia (HR 0.52 [95%CI 0.17-1.55]). Additionally, GLP-1RA use was associated with a significantly fewer hospitalizations (HR 0.74 [95%CI 0.62-0.90]) and ED (HR 0.73 [95%CI 0.57-0.92]).

### HTE Results

The details regarding data preprocessing and hyperparameter tuning were shown in **eResult 1 in the Supplement**. The estimated RD for DKA was negative in GLP-1RA users vs non-users (RD: -0.76% [95% CI: -1.44 to 1.29]), suggesting a potentially lower DKA risk associated with GLP-1RA but without statistical significance; the results were consistent with those from Cox regression models.

To further explore HTE, we employed interpretable decision tree models to identify patient subgroups with differential GLP-1RA effects on DKA (see **Figure 2**). Specifically, patients without hypothyroidism appeared to benefit from GLP-1RA treatment, particularly among those with dyslipidemia. In contrast, patients with hypothyroidism may experience potential harm (e.g., a positive risk difference).

**Figure 2.**
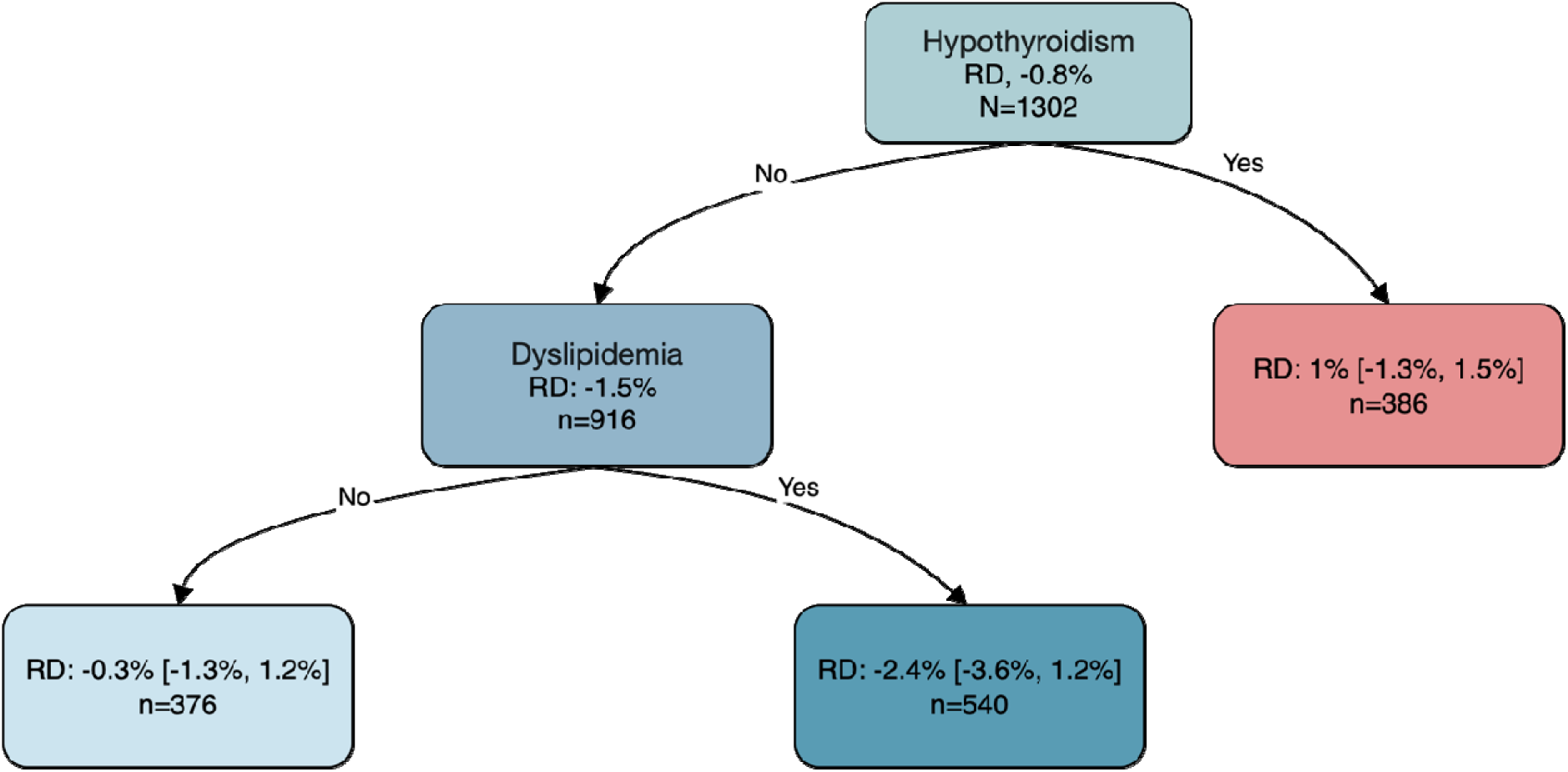
Interpretable decision tree for DKA.

### Effect modification assessment

There was a significant interaction between GLP-1RA and hypothyroidism (interaction HR, 1.48 [95% CI, 1.06–2.08]), suggesting that the benefit of GLP-1RA was attenuated among patients with thyroid conditions. Conversely, the interaction between GLP-1RA and dyslipidemia was not statistically significant (HR, 0.77 [95% CI, 0.56–1.08]), although a trend toward stronger benefit among dyslipidemia patients was observed.

A significant three-way interaction was found between GLP-1RA, hypothyroidism, and dyslipidemia (HR, 1.42 [95% CI, 1.04–1.94]), indicating that the combination of both conditions may further modify the treatment effect. These findings suggest that patients without hypothyroidism, particularly those with dyslipidemia, may derive greater benefit from GLP-1RA therapy in reducing the risk of DKA.

### Subgroup Analysis and Sensitivity analysis

The results for DKA are presented in **Figure 3**. The reduced risk of DKA associated with GLP-1RA use was particularly notable among individuals who had not used metformin at baseline (HR, 0.30 [95% CI, 0.10–0.91]). Additionally, we stratified the cohort based on potential moderators identified through interpretable decision tree analysis. GLP-1RA use was significantly associated with a reduced risk of DKA among patients without hypothyroidism (HR, 0.47 [95% CI, 0.24–0.92]), whereas no benefit was observed among those with hypothyroidism (HR, 1.08 [95% CI, 0.35–3.36]). Notably, the most pronounced benefit was observed among patients without hypothyroidism but with dyslipidemia compared to non-users (HR, 0.12 [95% CI, 0.03–0.55]).

**Figure 3.**
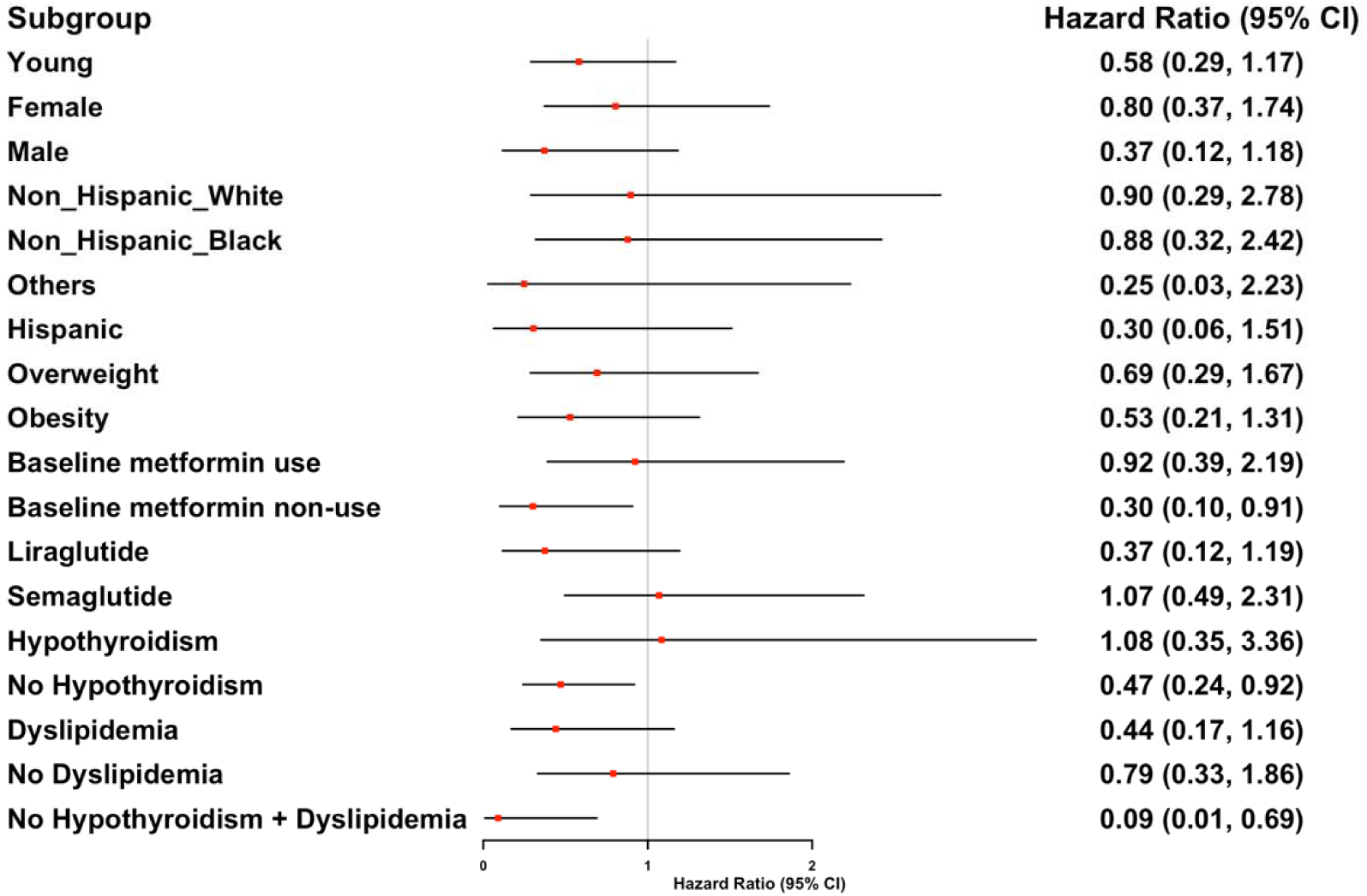
Subgroup analysis of DKA in patients receiving GLP-1 RA vs. those not receiving GLP-1 RA.

Regarding hospitalizations or ED visits (see **eFigure 4**), most subgroups demonstrated a significant reduction in risk associated with GLP-1RA use or showed a potential trend toward benefit.

Furthermore, we conducted a sensitivity analysis by composing DKA and severe hypoglycemia. While GLP-1RA use was not significantly associated with a reduced risk, it demonstrated a favorable trend (HR 0.65 [95% CI, 0.37–1.14]).

## Discussion

In our target trial emulation study leveraging real-world data from the OneFlorida+ EHR network, GLP-1RA use, compared to non-use, was not associated with a risk of DKA or severe hypoglycemia but linked fewer hospitalizations and ED visits among adults with T1D and obesity. While GLP-1RAs have consistently shown benefits in glycemic control, weight reduction, and cardiovascular outcomes among patients withT2D or obesity,^33,34^ evidence supporting their safe use in T1D remains limited and mixed, partly due to concerns about acute diabetes complications, such as hypoglycemia and DKA.^12,15,35^ Our findings contribute important real-world evidence to this understudied area, suggesting that GLP-1RAs may offer broader clinical benefits without introducing safety concerns on acute diabetes complications among individuals with T1D.

Prior studies evaluating GLP-1RA adjunct therapy in T1D have yielded mixed results. Early trials such as ADJUNCT ONE and TWO reported modest glycemic improvements but raised safety concerns, noting higher rates of hyperglycemia with ketosis with a high-dose GLP-1RA (liraglutide 1.8 mg).^35,36^ These findings tempered enthusiasm of GLP-1 RA use in T1D despite their marked benefits in improving cardiovascular and renal outcomes.^37,38^ In contrast, recent evidence suggests potential benefits of GLP-1RAs in T1D and Obesity.^39^ Meta-analyses and real-world studies indicate that adding a GLP-1RA can improve metabolic outcomes (lower HbA1c, weight loss, and reduced insulin requirements) without increasing the incidence of severe hypoglycemia or DKA.^40^ These prior observations set the stage for our investigation. In this context, our findings contrast with earlier safety concerns and instead underscore the safety of GLP-1RA regarding acute diabetes complications, as well as its broader clinical benefits in reducing hospitalizations and ER visits.

While our prespecified subgroup analyses were consistent with the main analysis overall, we observed that GLP-1RAs may offer the benefit of lowering DKA risk in particularly in patients who did not use metformin at baseline. One potential explanation is that both metformin and GLP-1RAs improve insulin sensitivity and reduce hepatic glucose output; their combined effects might lead to lower exogenous insulin requirements, which—if not carefully adjusted—can increase ketogenic risk.^41^ However, given the exploratory nature and limited power of these subgroup analyses, additional studies with larger sample size and longer follow-up are warranted to confirm these findings and to better characterize safety and effectiveness differences across subpopulations.

Using a data-driven approach, we identified that hypothyroidism may modify the association between GLP-1RA use and the DKA. Specifically, GLP-1RA use was associated with a lower risk of DKA among patients without hypothyroidism, but this protective association was not observed in those with hypothyroidism. Given that hypothyroidism is a common comorbidity in individuals with T1D—largely due to underlying autoimmune mechanisms—thyroid function may influence both glucose metabolism and the efficacy of adjunctive therapies such as GLP-1RAs.^42^ Hypothyroidism is linked to reduced insulin sensitivity, impaired glucose and ketone metabolism, and blunted counterregulatory responses, all of which may increase susceptibility to DKA.^43^ Also, both hypothyroidism and GLP-1RAs slow gastric motility, possibly affecting adherence and glucose control. These factors may attenuate the DKA risk reduction associated with GLP-1RAs observed in T1D patients without thyroid dysfunction.

Several limitations should be acknowledged when interpreting these results. First, despite the rigorous PS matching and adjustments for a broad range of covariates, residual confounding from unmeasured factors such as diet, physical activity, patient adherence, or socioeconomic status remains possible. In addition, the relatively limited number of events within specific subgroups led to wide confidence intervals. These results, therefore, must be interpreted cautiously, and future larger-scale studies or randomized controlled trials are needed to validate these findings.

## Conclusion

In conclusion, in this target trial emulation study of adults with T1D who had overweight or obesity, the use of GLP-1RAs was associated with fewer hospitalizations or ED visits, without increasing the risk of acute complications, including DKA and severe hypoglycemia. These findings provide valuable clinical evidence supporting the safety of GLP-1RAs use in this vulnerable patient population. Further research, particularly larger prospective studies and randomized controlled trials is warranted to confirm these findings.

## Author Contributions

Dr. J. Guo is the guarantor of this work and, as such, had full access to all the data in the study and takes responsibility for the integrity of the data and the accuracy of the data analysis.

Concept and design: A. Sheer, T. Hannon, M. Hayes, R. Sun, J. Guo, and J. Bian.

Acquisition, analysis, or interpretation of data: All authors.

Drafting of the manuscript: H. Dai, Y. Lee, and R. Radwan.

Critical review of the manuscript for important intellectual content: All authors.

Statistical analysis: H. Dai and Y. Lee

Obtained funding: J. Guo.

Supervision: J. Guo and J. Bian.

## Data Availability Statement

Data set Available through OneFlorida+ Clinical Research Network (email)

